# Longer incubation periods of SARS-CoV-2 infection in infants than children

**DOI:** 10.1101/2021.05.07.21256798

**Authors:** Char Leung

## Abstract

**Objective:** A large body of research has described the incubation period of SARS-CoV-2 infection, an important metric for assessing the risk of developing a disease as well as surveillance. While longer incubation periods for elderly have been found, it remains elusive whether this also holds true for infants and children, partly due to the lack of data. The present work clarified the incubation periods of COVID-19 for infants and children.

**Methods:** Using the data released by the Chinese health authorities and municipal offices, statistical comparisons of clinical features were made between infants (aged below 1 year) and children (aged between 1 and 17 years). An age-varying incubation period distribution period was modeled using maximum likelihood estimation modified for interval censored exposure time and age.

**Discussion:** Reported in 56 web pages, a total of 65 cases from 20 provinces dated between January and June 2020, including 18 infants and 47 children, were eligible for inclusion. Infants appeared to bear more severe clinical courses, as demonstrated by the higher prevalence of breathing difficulty as well as nasal congestion. In contrast, fever was less prominent in infants than in children. The incubation period was found to decrease with age, with infants appearing to have longer incubation periods.

**Conclusion:** Fever remained to be one of the most commonly seen symptoms in infants and children with SARS-CoV-2 infection and have continued to determine the time of symptom onset. While shorter incubation periods should be seen in patients with weaker immune system due to weaker antiviral response that is beneficial for viral growth, the longer incubation period in infants may be due to their weaker febrile response to the virus, leading to prolonged symptom onset.

## Introduction

Defined as the duration between exposure to the infectious source and the onset of symptoms, the incubation period is a measure of great public health and clinical importance. Since the outbreak in the end of 2019, a large amount of research has been devoted to measure the incubation period of COVID-19, a respiratory disease caused by SARS-CoV-2, using different sources of data, such as those available in news reports and press releases^1-3^, because epidemiologic data are usually rare. In addition, some studies have also investigated factors associated with the incubation period, including age^4^ and travel history^3^.

Age as a factor of incubation period of COVID-19 has previously been studied. In particular, it has been suggested that elderly tended to have longer incubation periods^5,6^. However, the incubation period for paediatric patients remains poorly known. In addition to age-specific quarantine policy making^4^, a better understanding of the incubation period also helps assessing the risk of developing COVID-19. Against this background, the present work aimed to clarify the difference in the incubation period between infants and children using the Chinese data.

## Materials and Methods

Because children are less affected by COVID-19, single centre studies on paediatric patients usually involve small samples hence fail to deliver informative results. The present work then follows existing methodologies using clinical data gathered from public sources ^1^, such as those reported by the media and local health authorities in China. The search strategy followed that previously described^3^. Multiple searches were performed on the Internet using the web browser Google for confirmed cases since the outbreak until 1^st^ July 2020 when daily new cases dropped below 100 per day. Cases reported after this date represented different clusters of cases. Search terms included “pneumonia” AND “novel coronavirus” AND “age” AND “discharge” in Chinese.

The following data were abstracted: (i) travel history to Hubei, (ii) gender, (iii) age, (iv) exact (or the range of) date of exposure to infectious agent, (v) date of first symptom onset, (vi) date of hospital admission, (vii) date of discharge, and (viii) symptoms on admission. The incubation period was calculated as the duration between exposure to the infectious agent and first symptom onset.

An individual case was included if all following inclusion criteria were met: (i) the patient was diagnosed with COVID-19, and (ii) the exact age of the patient is known and was below 18 years on admission. There was no language restriction.

The first part of the analysis concerned comparisons of baseline characteristics between infants and children. An infant was defined as a patient below a year of age whereas a child was defined as a patient aged between 1 and 17 years inclusively. Depending on the normality condition, t-tests or Mann-Whitney tests were performed on interval data, such as time from hospital admission to discharge, whereas Fishers exact tests were performed on categorial data, such as symptoms on admission.

In the second part, the distribution of the incubation period was estimated, assuming a lognormal distribution^1,7^ which was similar to Sartwell’s model^8^ consisting of a constant log-transformed median incubation period *µ* and dispersion factor *σ*. The median incubation period is the given by *e*^*µ*^. Here, however, the median incubation period is allowed to vary by age and is given by 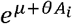, where *A*_*i*_ is the age of individual *i* where *θ* is the coefficient related to age. Therefore, a positive *θ* means that longer median incubation period is associated with older age.

Because the dataset contained both exact and interval-censored exposure time, the estimation method followed the maximum likelihood estimation (MLE) described in Reich et al^9^. For individual case, with exact date of exposure, *E*_*i*_, and symptom onset *S*_*i*_, the incubation time was defined as *E*_*i*_ − *S*_*i*_. The likelihood function for this individual case is given by 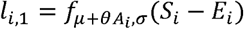, where *f* is the probability density function of lognormal distribution. If the date of exposure is not exactly known but believed to lie between *E*^*L*^ and *E*^*R*^, the likelihood function for this individual case is given by 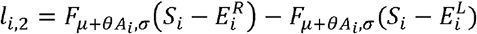, where *F* is the cumulative distribution function of lognormal distribution. Finally, parameters *µ, σ* and *θ* were estimated by maximising the sum of the loglikelihoods of all individual cases where each case is either *l*_*i*, 1_, or *l*_*i*, 2_.

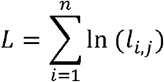

where *j* = 1, 2 This novel approach allows the use of all data in one estimation procedure. In contrast, estimating incubation period distributions by age groups results in smaller samples due to clustering.

All statistical tests and estimation were performed using the R software Version 3.6.1 and a p-value less than 5% was considered statistically significant.

## Results

Reported in 56 web pages, a total of 65 cases from 20 provinces dated between January and June 2020, including 18 infants and 47 children, were eligible for inclusion and the data were gathered. The two groups had similar share of male as well as travel history to Hubei, as shown in Table 1.

**Table 1.** Demographic and clinical characteristics of infants and children with COVID-19

|  | Infants<br>(n=18) | Children<br>(n=47) | p-value |
| --- | --- | --- | --- |
| Male, % | 50 (9/18) | 59.6 (28/47) | .579 |
| Travel to Hubei, % | 50 (9/18) | 40 (18/45) | .576 |
| Age, median, years | 0.4 (18) | 7 (47) | < .001 |
| Time from symptom onset to admission, median, days | 1.0 (14) | 1.0 (25) | .322 |
| Time from admission to discharge, median, days | 16.0 (15) | 16.0 (24) | .325 |
| Number of symptoms | 2.0 (16) | 1.0 (43) | .192 |
| Symptoms, % |  |  |  |
| Cough | 50.0 (8/16) | 20.9 (9/43) | .050 |
| Dyspnoea / breathing difficulty / chest discomfort | 23.5 (4/17) | 2.3 (1/43) | .020 |
| Fever | 31.2 (5/16) | 68.9 (31/45) | .016 |
| Lethargy | 6.2 (1/16) | 2.3 (1/43) | .472 |
| Vomiting/ nausea | 0 (0/16) | 4.7 (2/43) | > .999 |
| General pain | 0 (0/16) | 4.4 (2/45) | > .999 |
| Diarrhoea | 0 (0/16) | 4.7 (2/43) | > .999 |
| Sore throat | 0 (0/16) | 11.1 (5/45) | .313 |
| Nasal congestion/<br>rhinorrhoea | 29.4 (5/17) | 2.3 (1/44) | .005 |

Two measures concerning hospital admission including time from symptom onset to admission and time from admission to discharge saw no significant difference between the two groups. Infants appeared to have more symptoms compared with children, yet the difference was not significant.

Fever and cough remained the most commonly seen symptoms in both groups, like in adults. Vomiting/ nausea, general pain, and diarrhoea were not observed in infants. However, it is important to note that the absence of some of these symptoms in infants such as general pain might be due to the lack of verbal skills of infants. Nevertheless, there are signs infants bearing more severe clinical courses, compared with children. Infants had ten-fold (23.5% vs 2.3%) and significantly higher prevalence of dyspnoea / breathing difficulty / chest discomfort. Moreover, nasal congestion / rhinorrhoea was more commonly seen in infants. Interestingly, fever was less prominent in infants (31.3% vs 68.9%). Overall, infants appeared to be more vulnerable to both lower and upper respiratory tract infections than children^10^.

The estimates of the incubation period distribution are shown in Table 2. All estimates are significant at 5%. The coefficient for age *θ* is negative, meaning that shorter incubation period is associated with increased age. Figure 1 visually illustrates the relationship between age and the median incubation period using the estimates reported in Table 2. The model suggests that the median incubation period is about 5.8 days in newborns, as implied by Figure 1.

**Table 2.** Estimates of the incubation period distribution

|  | Estimate |
| --- | --- |
| $\mu$ | 1.749** |
| $\sigma$ | 0.637** |
| $\theta$ | -0.046* |
| AIC | 69.913 |

Significant at \*5% and \*\*1%

**Figure 1.**
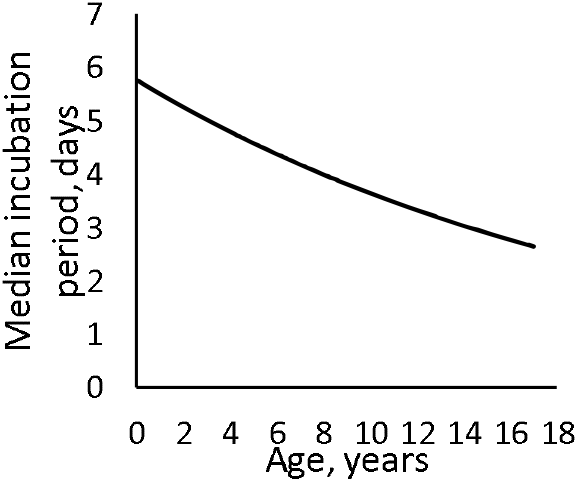
Median incubation period associated with age

## Discussion

The present work showed that children was associated with shorter incubation period, compared with infants. While reduced severity in paediatric patients has been commonly observed^11^, infants appeared to be more susceptible to lower respiratory tract infection.

While some studies have investigated the age-varying incubation period of COVID-19^4-6^, little attention has been paid to paediatric patients. Xiao et al^12^ has proposed a U-shaped relationship between age and incubation period. The authors estimated confidence intervals for the median incubation periods by age groups. However, sample stratification by age reduces the sample size of each age group, leading to wide confidence intervals and failing to produce conclusive results. In contrast, the novel approach proposed in the present work models the age-dependent incubation period without stratification by age that reduces the sample size.

It is generally believed that the incubation period depends on the initial infective dose, and the speed of the replication of the pathogen and defence mechanisms within the host^13^. Although it follows that patients with weaker immune system should see shorter incubation periods because weaker antiviral response beneficial for viral replication, it has also been suggested that the more responsive the immune system is to the virus, the shorter the incubation period^14^.

The relationship between the incubation period and immune system (and disease severity) also depends on the types of symptoms associated with the disease because the incubation period takes into account the symptom onset. For respiratory virus infection, most symptoms are results of immune responses to the infection while some are directly related to the viral cytopathic effect, such as shedding of damaged epithelium leading to airway obstruction^15^. For COVID-19, fever is the most commonly observed symptom with prevalence of at least 80%^16-18^. It is an immune response to the pathogen and is induced by pyrogenic cytokines such as IL-1, IFN and IL-6^19,20^. In infants, fever may be less marked due to weaker immune response, as evidenced by low IFN-γ level^21,22^. Consequently, infants as well as patients with less responsive immune system should see longer incubation periods^14^. Similarly, there has been evidence suggesting longer incubation period in elderly^5,12,23^ and immunocompromised patients^24,25^ with COVID-19. In particular, it has been suggested that the longer incubation period in the elderly was due to the weaker febrile response^23^.

While infants appeared to have higher prevalence of nasal congestion / rhinorrhoea as well as dyspnoea / breathing difficulty / chest discomfort, these symptoms are less likely to be associated with the longer incubation periods observed in neonates because they are generally less prominent in COVID-19 patients, compared with fever. In addition, dyspnoea typically develops a few days after the onset of other symptoms^26^, making fever the hallmark of first symptom.

Selection bias is the major limitation of the present work. Although the choice of the search terms inevitably leads to bias, data of incubation period are relatively rare thus the public domain remains an alternative source of information, as demonstrated in existing literature^1-3^.

## Conclusion

The present work clarified the age-varying incubation period of SARS-CoV-2 infection among different paediatric groups, the first study of its kind. Fever remained to be the most commonly seen symptom in infants and children with SARS-CoV-2 infection and have continued to determine the time of symptom onset. While shorter incubation periods should, by theory, be seen in patients with weaker immune system due to weaker antiviral response that is beneficial for viral growth, the longer incubation period in infants was due to their weaker febrile response to the virus, leading to prolonged symptom onset.

## Data Availability

Data are available upon reasonable requests

## Abbreviation

PCR: polymerase chain reaction

